# A new fMRI localizer for preoperative language mapping using a sentence completion task: Validity, choice of baseline condition, and test-retest reliability

**DOI:** 10.1101/2021.10.17.21264884

**Authors:** Kirill Elin, Svetlana Malyutina, Oleg Bronov, Ekaterina Stupina, Aleksei Marinets, Olga Dragoy

## Abstract

To avoid post-neurosurgical language deficits, intraoperative mapping of the language function in the brain can be complemented with preoperative mapping with fMRI. The validity of an fMRI ‘language localizer’ paradigm crucially depends on the choice of an optimal language task and baseline condition. This study presents a new fMRI ‘language localizer’ in Russian using overt sentence completion, a task that comprehensively engages the language function by involving both production and comprehension at the word and sentence level. The paradigm was validated in 18 neurologically healthy volunteers who participated in two scanning sessions, for estimating test-retest reliability. For the first time, two baseline conditions for the sentence completion task were compared.

At the group level, the paradigm significantly activated both anterior and posterior language-related regions. Individual-level analysis showed that activation was elicited most consistently in the inferior frontal regions, followed by posterior temporal regions and the angular gyrus. Test-retest reliability of activation location, as measured by Dice coefficients, was moderate and thus comparable to previous studies. Test-retest reliability was higher in the frontal than temporo-parietal region and with the most liberal statistical thresholding compared to two more conservative thresholding methods. Lateralization indices were expectedly left-hemispheric, with greater lateralization in the frontal than temporo-parietal region, and showed moderate test-retest reliability. Finally, the pseudoword baseline elicited more extensive and more reliable activation, although the syllable baseline appears more feasible for future clinical use.

Overall, the study demonstrated the validity and reliability of the sentence completion task for mapping the language function in the brain. The paradigm needs further validation in a clinical sample of neurosurgical patients. Additionally, the study contributes to general evidence on test-retest reliability of fMRI

## 1 Introduction

### 1.1 Language mapping in neurosurgical patients

When patients undergo neurosurgical interventions for brain tumors, refractory epilepsy, arteriovenous malformations et cetera, a crucial goal is to remove pathological tissue while sparing eloquent (functionally necessary) areas, so that the respective functions, including cognitive ones (Satoer et al., 2016), are not impaired following neurosurgery (Duffau, 2012). One critical function is language processing: it lies at the core of human communication, and its impairment negatively impacts return to work, social inclusion, and general quality of life (Gabel et al., 2019, Hilari et al., 2003).

Localization of the language network is highly variable across individuals (Ojemann, 1979), and the variability is further enhanced by functional re-organization that happens in case of brain pathology: for example, over the course of brain tumor growth (Almairac et al., 2018, Zhang et al., 2018). Thus, to avoid damage to brain areas critical for the language function and to prevent subsequent language impairment, the neurosurgical team performs mapping of ‘language-eloquent’ brain areas in individual patients. The gold standard for localizing language-eloquent brain areas is intraoperative mapping with direct electrical stimulation (DES) during awake craniotomy (Ojemann, 1979, Rofes et al., 2019). During this procedure, an electric current is applied to exposed brain tissue, causing a temporary disruption of neural activity. Meanwhile, the patient is awake from anaesthesia and is performing a language task. If application of DES to an area reliably leads to errors or speech arrest, this means that the area is eloquent and should be spared during neurosurgery, if possible.

While intraoperative DES is the standard procedure for language mapping, there are reasons to complement it with additional preoperative mapping. Firstly, preoperative language mapping allows to plan the surgical procedure in advance (Silva et al., 2018, Weng et al., 2018). Based on preoperative mapping data, the neurosurgeon can decide whether intraoperative DES mapping of the language function is necessary (for example, when operating over a presumably non-language-dominant hemisphere) or plan an optimal access route to bypass language-eloquent areas. Secondly, data from preoperative mapping can be used if DES cannot be completed: for example, if the patient does not cooperate or if there are epileptic seizures during DES (Hervey-Jumper et al., 2015). In such cases, the results of preoperative mapping, even though providing less direct information than DES, can still inform the neurosurgeon and reduce the risks of functional impairment.

Historically, preoperative language mapping was performed using the intracarotid sodium amobarbital procedure, known as the Wada test (Wada & Rasmussen, 1960). The test involves anesthetizing one hemisphere by injecting sodium amobarbital through a catheter while the patient is performing a language task. Failure to perform the task indicates that the anaesthesized hemisphere is crucial for language processing. However, the Wada test has major limitations. It is a highly invasive procedure that can cause complications: according to Loddenkemper, Morris and Möddel (2008), they emerge in up to 11% patients and include serious adverse events such as stroke. Another limitation is that effects of sodium amobarbital only last for a few minutes, providing very limited time for testing (Loring, Meador, & Lee, 2002). Critically, the Wada test can only assess hemispheric dominance of language processing but cannot address specific localization of language-eloquent areas within a hemisphere.

Therefore, other technologies have been replacing the Wada test for preoperative language mapping: navigated transcranial magnetic stimulation (nTMS; Picht et al., 2013), functional magnetic resonance imaging (fMRI; for review, see Agarwal et al., 2019, Silva et al., 2018), magnetoencephalography (MEG; Van Poppel et al., 2012), or combination thereof (Ille et al., 2015, Sollman et al., 2016). Unlike the Wada test, these methods are largely safe and non-invasive. On top of that, they have high spatial resolution and allow to identify language-eloquent brain areas with the precision of millimeters. To take full advantage of these methods, it is crucial to choose an optimal functional paradigm that would be sensitive, specific and reliable in identifying both lateralization (hemispheric dominance) and specific localization of brain networks comprehensively enabling the language function. In this paper, we present such paradigm for pre-operative language mapping using fMRI in Russian-speaking individuals and provide methodological evidence on its test-retest reliability and the optimal baseline condition.

### 1.2 Choice of a language task for fMRI mapping

The quality of a language mapping paradigm critically depends on the choice of a language task: that is, whether it is able to comprehensively engage all levels of linguistic processing while remaining feasible. Previous fMRI ‘language localizer’ paradigms have used a variety of tasks (for review, see Bradshaw et al., 2016, Manan et al., 2020). Below, we review most popular language tasks and summarize previous evidence on their success in identifying language networks in the brain.

Most ‘language localizers’ have used single-word tasks, particularly expressive single-word tasks (Manan et al., 2020). A traditional expressive single-word task adapted from early intraoperative batteries (Ojemann, 1993, Ruge et al., 1999) was number counting. However, counting is a highly automated process that engages linguistic processing only superficially and is no longer considered sufficiently sensitive to identify language networks (Morrison et al., 2016b; Petrovich Brennan et al., 2007). Today, other tasks are used to engage word retrieval, such as picture naming (Petrovich Brennan et al., 2007, Pouratian et al., 2002, Roux et al., 2003, Rutten et al., 2002) and verbal fluency, where the participant has to name as many words as possible from a given semantic category or starting with a given letter (Ruff et al., 2008 Sanjuan et al., 2010). Expressive single-word tasks are intuitive for the participant and are easily timed relative to fMRI scanning. Still, they only engage the sound and word levels of language production, leaving out any grammatical processing and any language comprehension. Thus, they cannot fully identify brain areas that are crucial for sentence-level communication. Indeed, compared to sentence-level tasks, expressive single-word tasks elicit less activation in both anterior and posterior left-hemispheric ‘language regions’ (Połczyńska et al., 2017, Manan et al., 2020). Additionally, picture naming shows a poor lateralizing ability (Bradshaw et al., 2017, Deblaere et al., 2002).

Similar problems are faced by receptive single-word tasks, such as phonemic judgement, requiring to make a decision about the sound structure of a word (for example, whether two words rhyme; Jones, Mahmoud & Philipps, 2011), or semantic judgement, requiring to make a decision about the meaning of a word (for example, whether it refers to an animate object, or whether two words are opposite in meaning; Binder et al., 1996, Szaflarski et al., 2008). Again, such tasks are easily integrated with the timing of fMRI scanning. Moreover, unlike expressive single-word tasks, they do not evoke head motion artifacts due to articulation. However, they are even more limited in engaging linguistic processes and cannot detect brain networks enabling language production or any grammatical processing beyond the word level. Empirically, these tasks have shown limited lateralizing ability (Deblaere et al., 2002, Jansen et al., 2006) and reliability (Jansen et al., 2006; reliability will be discussed in more detail below).

To more fully activate language networks, a number of fMRI protocols used sentence-level tasks requiring to process not only individual words but also their grammatical and semantic relations. Examples of expressive sentence-level tasks are describing a picture with a sentence (Mauler et al., 2017, Partovi et al., 2012) or generating a sentence with given words (Hakyemez et al., 2016). Such tasks appear to successfully activate both anterior and posterior language areas (Mauler et al., 2017, Partovi et al., 2012, Hakyemez et al., 2016). However, they are taxing for the patient and difficult to time relative to fMRI scanning, especially given interindividual variability in task completion speed across patients, so they are not widely used.

Much more popular are receptive sentence-level tasks. These are sentence or passage listening (Pillai & Zaca, 2011, Suarez et al., 2014, Wilson et al., 2017) or reading (Grummig et al., 2006, Fedorenko et al., 2010), which may be passive or accompanied by comprehension questions, such as to judge real-world plausibility of a sentence or match it to a picture (Kinno et al., 2014, Pillai & Zaca, 2011). These tasks are more easily timed than expressive sentence-level tasks but also successfully engage grammatical processing: that is, the participant has both to process individual words and analyse their relations. Passive receptive sentence-level tasks have an additional advantage of feasibility in patients with compromised language production or non-cooperative patients (for example, in the pediatric population, Suarez et al., 2014) but also an additional drawback: it is impossible to control or even monitor how much the participant is engaged in the task. Crucially, a major limitation is that none of receptive sentence-level tasks engage brain networks crucial for language production. Empirically, these tasks have shown low lateralizing abilities (Lehéricy, 2000; Pillai & Zaca, 2011).

Taken together, in order to comprehensively identify brain networks that enable real-life language use, an fMRI paradigm needs to engage both language production and comprehension in a task that goes beyond the word level. One solution is a conjunction analysis of multiple tasks targeting different language processes separately (Pouratian et al., 2002, De Guibert et al., 2010). However, interpretation of the conjunction analysis is not straightforward if tasks elicit largely different activations. Additionally, from the clinical viewpoint, multiple-task paradigms are time-consuming and less feasible in clinical settings. Thus, another solution is an fMRI localizer paradigm that uses a single task engaging both language production and comprehension beyond the word level.

One such comprehensive task is sentence completion, advocated in a recent white paper of the American Society of Functional Neuroradiology (Black et al., 2017) and a metaanalysis by Manan et al. (2020). In this task, the participant has to read aloud a sentence with a missing final word and complete it with a semantically and grammatically appropriate word. This task comprehensively involves many linguistic processes in both comprehension (orthographic processing, word access, grammatical parsing, semantic integration) and production (word search, grammatical inflection in morphologically complex languages such as Russian, phonological encoding and articulation). Empirically, previous works proved sentence completion superior to other tasks in assessing both lateralization and localization of language processing networks (Salek et al., 2017, Połczyńska et al., 2017, Zacà et al., 2012, Barnett et al., 2014, Wilson et al., 2017, Unadkat et al., 2019).

Inspired by these sentence completion paradigms in English, the present study presents a similar paradigm in Russian. Russian is the 8th most spoken language in the world, with about 120 million first-language speakers worldwide (Eberhard, Simons & Fennig, 2020), so a new clinical tool in Russian would serve the needs of a large Russian-speaking clinical population. So far, Russian-language paradigms for presurgical language mapping have been very few and have never used a sentence completion task (Litvinova et al., 2012, Rumshiskaya et al., 2014).

### 1.3 Choice of a baseline task for fMRI mapping

A crucial concept in classic fMRI analysis is ‘subtraction logic’: to isolate neural activation related to the process of interest, the analysis should ‘subtract’ the activation in a ‘lower-level’ baseline (control) task from the activation in a ‘higher-level’ experimental task (Huettel et al., 2008). Specifically in case of ‘language localizer’ paradigms, such subtraction allows to isolate language-related neural activity from activity due to sensorimotor processes, general alertness, et cetera. Due to the subtraction principle, not only the choice of an experimental language task but also the choice of a lower-level baseline task can vastly impact the findings of fMRI ‘language localizers’ (Bradshaw et al., 2017).

Some previous fMRI ‘language localizers’ used passive rest or viewing of a fixation cross as the baseline condition (Jones et al., 2011, Suarez et al., 2014). However, a passive baseline is problematic for several reasons. Firstly, a passive baseline does not require any sensorimotor or cognitive activity, so subtracting it from the experimental condition does not fully isolate language-related activity from lower-level processes. Secondly, it is not possible to control or monitor the patient’s cognitive activity during passive rest, so mind-wandering or other patient-initiated cognitive activity may confound the results. Indeed, fMRI analyses using passive baselines have elicited less specific and less lateralized activation compared to analyses using active baselines (Dodoo-Schittko et al., 2012, Hund-Georgiadis et al., 2001, although see Miró et al., 2014, Newman et al., 2001).

Therefore, active baselines are typically recommended for more specific isolation of language-related activity from lower-level processes (Bradshaw et al., 2017). Choosing an optimal active baseline presents a challenge: for most language tasks, the respective lower-level processes can be addressed by several theoretically possible baselines. For example, for listening tasks, the baseline condition can involve listening to backwards speech (Lehéricy et al., 2000, Thivard et al., 2005), various types of noise (Rodd et al., 2005), or music (Bleich-Cohen et al., 2009). Although all these baselines involve auditory processing (lower-level sensory processing) and presumably no linguistic processing, an empirical comparison by Stoppelman et al. (2013) showed that different baselines yielded very different results. So far, such direct empirical comparisons in order to choose an optimal baseline have only been made for few language tasks and baseline types (Binder et al., 2008, Newman et al., 2001, Stoppelman et al., 2003).

To the best of our knowledge, our study is the first to make an empirical comparison between two different baselines for the sentence completion task. These are a syllable baseline, where the participant has to read aloud a sequence consisting of the same syllable and repeat the syllable once more, and a pseudoword baseline, where the participant has to read aloud a sequence of pseudowords and repeat any of them once. Both baselines are theoretically plausible. In contrast to the experimental condition, they do not consist of real words or resemble grammatical structures, so they do not elicit any linguistic processing. At the same time, they involve the same ‘lower-level’ processes as the experimental condition: visual and orthographic processing, motor planning and articulation, and initiation of a response (completion of a sequence). Thus, their subtraction allows to maximally isolate language-related from ‘lower-level’ neural activity. Previous sentence completion paradigms used other baselines that subtracted ‘lower-level’ activity less fully (rest: Połczyńska et al., 2017, passive viewing of nonsense symbols: Barnett et al., 2014, Zacà et al., 2012) or a conjunction analysis approach without an explicit baseline (Wilson et al., 2017). The present study is the first to employ and compare a syllable and pseudoword baseline for the sentence completion task.

### 1.4 Reliability of fMRI mapping

Another methodological contribution of the present study is estimating test-retest reliability of the fMRI paradigm. Reliability is critical for any clinical usage of fMRI ‘language localizers’: distribution of brain activity needs to be reproducible at multiple testing sessions in order to consider it clinically meaningful and draw any implications for neurosurgical treatment. Recent studies have raised concerns about test-retest reliability of task-based fMRI in general, due to inherent physiological noise, scanner noise, changes in concurrent non-task-related cognitive activity in participants, et cetera (Bennett & Miller, 2010, Elliott et al., 2020, Holiga et al., 2018). In light of these general concerns, it is important to quantify and report reliability of any paradigms suggested for clinical use.

Previous studies have started estimating test-retest reliability of fMRI ‘language localizer’ paradigms in healthy control participants (Fesl et al., 2010, Morrison et al., 2016a, Nettekoven et al., 2018, Wilson et al., 2016) and clinical populations (Fernández et al., 2003, Morrison et al., 2016a). For example, Morrison et al. (2016a) showed high individual variability in test-retest reliability, which was on average lower in a phonemic fluency task than in a rhyming task, and in patients with high-grade gliomas than patients with low-grade gliomas and healthy control participants. Using an overt object naming task in healthy participants, Nettekoven et al. (2018) showed high reliability of the activation peak location but low reliability of activation extent, particularly in the right hemisphere.

To the best of our knowledge, only two studies so far have estimated test-retest reliability of sentence completion paradigms. Whalley et al. (2009) used the Hayling sentence completion task in individuals with high genetic risk of schizophrenia and showed good test-retest reliability. Wilson et al. (2016) compared test-retest reliability of four language tasks in healthy participants and found the best reliability in picture naming, followed by naturalistic comprehension, sentence completion, and narrative comprehension. Despite this pattern, the authors concluded that sentence completion was one of two tasks offering the best balance of reliability and validity for an fMRI language localizer. Our study aims to add to these emerging data and provide more evidence on test-retest reliability of a sentence completion fMRI paradigm.

Previous studies used different metrics to quantify test-retest reliability: Dice coefficient (Fesl et al., 2010, Morrison et al., 2016a, Nettekoven et al., 2018, Wilson et al., 2016), Jaccard index (Morrison et al., 2016a), Euclidean distance (Morrison et al., 2016a, Nettekoven et al., 2018), voxelwise intraclass correlation coefficient (Fernández et al., 2003, Nettekoven et al., 2018, Whalley et al., 2009), correlation of lateralization indices (LIs; Morrison et al., 2016a, Fesl et al., 2010). The present study adopted two of them: between-session Dice coefficient and correlation of lateralization indices. Advantages of the Dice coefficient are that it is widely used in the literature, is straightforward to interpret and, unlike intraclass correlation coefficient, can provide a global whole-brain measure and is calculated individually with no reference to group data (Bennett & Miller, 2010, Wilson et al., 2016). Besides, using a Dice coefficient ensures comparability to Wilson et al. (2016), the only previous study that included a sentence completion task and compared its reliability to other tasks. Additionally, we measured the test-retest correlation of LIs because, just as individual activation maps, they also present a clinically relevant measure that can inform a neurosurgeon’s decision on the necessity of awake surgery.

### 1.5 The present study

To summarize, this paper presents a new fMRI localizer paradigm for preoperative language mapping in Russian-speaking individuals with brain tumors, refractory epilepsy, and other conditions when neurosurgery is indicated. Following the best practices in other languages, we used a sentence completion task that comprehensively engages language production and comprehension processes at the word and sentence level. We present the data from a control group of neurologically healthy individuals, test whether the paradigm can successfully identify the expected key language-related areas in this group, compare two different baseline conditions (syllables versus pseudowords), and quantify the test-retest reliability of the paradigm.

## 2 Method

### 2.1 Participants

The study included 21 right-handed native speakers of Russian with no history of neurological or psychiatric disorders. Data of three participants were excluded from analysis due to excessive head movement in the scanner (more than 5 mm), resulting in a sample of 18 participants (14 females; age: mean 41.3, SD 6.6, range 30–53 years; years of education: mean 16.2, SD 4.7, range 11–30 years; Edinburgh Handedness Inventory score: mean 52, SD 2.67, range 46–55). All participants had normal hearing and normal or corrected-to-normal vision. All participants gave written informed consent.

### 2.2 Task and Stimuli

During each scanning session, participants performed two identically structured language mapping paradigms. Both mapping paradigms comprised an experimental condition and a baseline condition. The experimental condition was identical in the two paradigms: participants were visually presented with a Russian sentence with a missing final word and instructed to read the sentence aloud and produce an appropriate final word aloud. The two paradigms differed with regard to the baseline condition, including either a syllable (henceforth, SYLL) or a pseudoword (henceforth, PW) baseline. In the SYLL baseline condition, participants read aloud a visually presented string consisting of one syllable repeated three times (e.g., «*Φеее фееееее фееееее* …*»* - «*Feee feeeeee feeeeee* …») and repeated the syllable aloud one more time. In the PW baseline condition, participants read aloud a visually presented string of pseudowords (pseudonouns) phonotactically legal in Russian (e.g., «*Уптилья пикаш измеха* …» - «*Uptilja pikaš izmeha* …») and repeated any single of the pseudowords.

The stimuli in the experimental condition were Russian sentences (60 per paradigm). The full stimuli list is publicly available online: https://www.hse.ru/en/neuroling/research/fmri-mapping. The sentences were three words long and had one of the following syntactic structures:

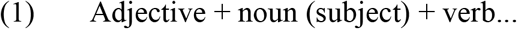

*Умная соседка прочла …*.

*A clever neighbour read…*

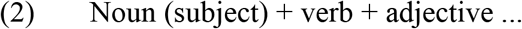

*Cкрипачка сдала сложный …*

*A violinist passed a challenging …*

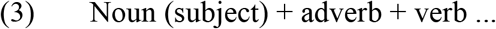

*Грабитель ловко украл …*

*A thief skillfully stole…*

All verbs were used in the present or past tense and required a direct object. In sentences of structure (2), the inflectional form of the adjective unambiguously determined the gender and number of the direct object. Words were no longer than three syllables and at least one word in each sentence was no longer than two syllables, resulting in the mean length of 7.38 syllables per sentence (SD .75, range 5–8). In both paradigms, verb tense and subject gender could repeat in no more than two consecutive trials both within and across presentation blocks consisting of three sentences (see 2.4. Procedure), with one exception of three consecutive past tense forms per paradigm.

The sentences were selected from a set of 160 sentences tested in an online pilot study, where 100 participants (50 females, age: mean 38.3, SD 11.5, range 18–68 years) read the sentences and finished them aloud with a semantically plausible word within five seconds, matching the task timing in the fMRI study. Their responses were recorded and scored for accuracy by a single rater. A response was considered accurate if it was a semantically plausible single word in a grammatically correct form (direct object). Based on the results, 120 sentences were selected and split into two halves with similar accuracy, to be used in the SYLL and PW paradigm (SYLL: mean accuracy 90.8%, SD 5.8%, range 78% – 100%; PW: mean accuracy 89.9%, SD 5.4%, range 80% – 100%; *t*(118) = .842, *p* = .401).

The two final lists were matched for the gender of the subject (31 feminine, 29 masculine), sentence structure (type 1: n=20, type 2: n=19, type 3: n=21), number of present and past verb forms (SYLL: 30 present / 30 past forms, PW: 29 present / 31 past forms), length in syllables (SYLL: mean 7.43, SD .76, range 5–8; PW: mean 7.31, SD .74, range 5–8; *t*(118) = .843, *p* = .400) and overall word frequency (SYLL: mean 39.37, SD 55.18, range .4–424.1; PW: mean 33.75, SD 45.71, range .5 – 277; *t*(358) = 1.051, *p* = .293).

In the SYLL baseline condition, stimuli were 60 phonologically legal Russian syllables (consonant + vowel), where the vowel was spelled multiple times in order to match the experimental condition for length in letters (for example, for the syllable “*фе*” - “*fe*”, the stimulus was «*Феее фееееее фееееее* …» - «*Feee feeeeee feeeeee* …»). Participants were instructed to ignore the exact number of vowel letters and pronounce the syllable duration approximately. In the PW baseline condition, the stimuli were 60 pseudowords phonotactically legal in Russian, constructed to pairwise match the number of syllables in experimental sentences. All pseudowords were constructed as pseudonouns: that is, none of them had inflection typical for other parts of speech.

### 2.3 MRI data acquisition

MRI data were obtained on a Siemens Magnetom Skyra 3T scanner with a 20-channel head coil at the National Medical and Surgical Center named after N.I. Pirogov of the Ministry of Healthcare of the Russian Federation. Participants wore MRI-compatible headphones to reduce scanner noise and head movement. Visual stimuli were presented using head-coil mounted goggles (NordicNeuroLab, Bergen, Norway). Stimuli presentation was controlled with nordicAktiva, version 1.2.1. Oral responses were recorded with an MRI-compatible FOMRI III™ microphone (Optoacoustics LTD) using OptiMRI recording software, version 3.1.

For anatomical reference, T1-weighted MPRAGE structural images were acquired with the following parameters: voxel size 1.0×1.0×1.0 mm, 176 axial slices in ascending order, slice thickness 1.00 mm, field of view (FoV) 320×320 mm, repetition time (TR) 2200 ms, echo time (TE) 2.43 ms, flip angle 8°. Functional blood-oxygenation-level-dependent (BOLD) data were obtained using the following parameters: voxel size 3.0×3.0×3.0 mm, 30 oblique slices in interleaved order, slice thickness 3 mm, FoV 205×205 mm, TR 7000 ms, TE 30 ms, delay in TR 5000 ms (sparse sampling), flip angle 90°, 128 volumes per paradigm.

### 2.4 Procedure

Each participant was scanned on two separate occasions with an interval of 14 days. The first session began with a short instruction and practice to familiarize participants with the task outside the scanner. Inside the scanner, acquisition of T1 anatomical images was followed by the two functional paradigms (SYLL and PW). Their order was balanced across participants and remained constant in the two sessions.

Each paradigm started with visually presented instructions followed by one training block per condition (not analyzed). Then, 120 stimuli (60 experimental and 60 baseline) were presented in blocks of three (Figure 1). A sparse sampling procedure was used (Hall et al., 1999). During a five-second delay in TR, participants gave oral response to the current stimulus (Figure 1). They were instructed to remain silent during the next two seconds, indicated by a «!» sign. During this time, MR images were acquired. The sparse sampling procedure allowed to minimize motion-induced artifacts due to articulation and to monitor participants’ responses with no acoustic noise from scanning. The duration of each paradigm was 14 min 56 sec in total.

**Figure 1.**
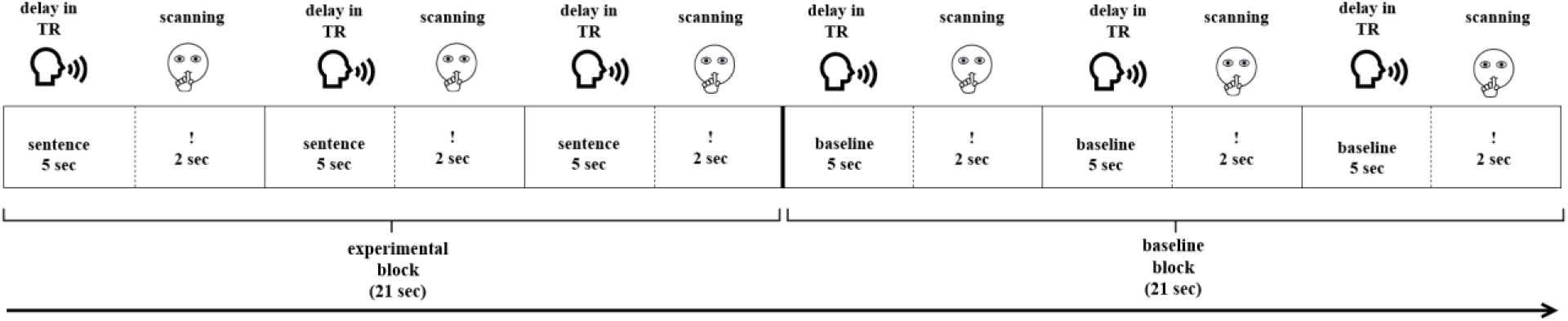
Experimental design of the functional paradigms.

### 2.5 Data analysis

#### 2.5.1 Behavioral data

The participants’ auditory responses from both sessions were transcribed, except for one participant’s responses that were lost due to technical error. Response accuracy was assessed independently by two raters. In the experimental condition, a response was considered accurate if it was a grammatically correct and semantically appropriate sentence completion. In the baseline conditions, a response was considered accurate if the participant read and repeated a syllable/pseudoword without any phonological errors. Inter-rater reliability, as assessed using percent agreement and Cohen’s kappa (Cohen, 1960), was high (1^st^ session: 98.36% and .76, respectively; 2^nd^ session: 98.62% and .69, respectively). All inconsistencies were resolved by discussion between the two raters.

#### 2.5.2 Activation maps

MRI data were analyzed using SPM12 (http://www.fil.ion.ucl.ac.uk/spm/software/spm12/) for MATLAB 2014b. Prior to data analysis, first eight volumes of each functional paradigm, corresponding to instructions and training blocks, were discarded. For data preprocessing, images of each participant were first manually reoriented to the AC-PC plane. Then, functional scans were re-aligned to correct for head motion. Participants with excessive head movement (more than 5 mm in any direction) were excluded from further analysis. Functional images were coregistered to the anatomical T1 image, followed by spatial normalization of images to the International Consortium of Brain Mapping (ICBM) space template–European brains (Mazziotta et al., 1995) based on segmentation into six tissue types (grey matter, white matter, cerebrospinal fluid, bone, soft tissue and air/background) defined by tissue probability maps in SPM12. This step was followed by spatial smoothing with an isotropic 8-mm Gaussian kernel.

Statistical analysis was performed separately for each paradigm and each session, resulting in four activation maps for each participant. In the first-level (individual-level) analysis, a high-pass filter with a cut-off period of 256 s was employed to remove slow signal drift. The model included two conditions: experimental (sentence completion) and baseline (SYLL or PW, depending on the paradigm). The duration of each event was set to 7 s. Six movement parameters obtained in re-alignment were entered as regressors. A canonical hemodynamic response function with no derivatives was used to model BOLD response. Model estimation was done using a restricted maximum likelihood fit. T-contrast maps were computed separately for each paradigm, subtracting activation in the baseline condition (SYLL or PW) from activation in the experimental condition.

Although the present paper focuses on individual localization of language-related areas, we still conducted second-level (group) statistical analysis for illustrative purposes, based on activation maps from the first session. The contrast maps from the first-level analyses were submitted to the second-level one-sample t-test. To visualize the results at different levels of statistical stringency, three types of statistical thresholding were applied to all group fMRI activation maps: the most conservative Bonferroni correction for multiple comparisons (family-wise error correction, FWE) at *α* = .05; a more liberal cluster-size correction with a minimum cluster size of *k* ≥ 200 mm3 at *α* = .001; and adaptive thresholding (AT) method proposed by Gorgolewski et al. (2012), which combines Gamma-Gaussian mixture modeling with topological FDR thresholding at *α* = .05. The AT method takes into account the strength of the signal: it generates lower thresholds when the signal is weak, resulting in fewer false negative clusters, and higher thresholds when the signal is strong, resulting in fewer false positive clusters, aiming to ensure an optimal balance between Type I and Type II error rate (Gorgolewski et al., 2012).

Anatomical labels for activation clusters were determined based on the Brainnetome atlas (Fan et al., 2016) implemented in the ICN_atlas toolbox (Kozák et al., 2017) for SPM12. The Brainnetome atlas was selected due to detailed parcellation, as it includes 246 regions in total. The activation maps were visualized in MRIcroGL 1.2.2 (https://www.nitrc.org/projects/mricrogl).

#### 2.5.3 Assessing individual-level activation of ‘key language-related areas’

Since the ultimate goal of the localizer was to individually localize critical language areas, we estimated how well the paradigms were able to activate them in each participant. Following Benjamin et al. (2017), we focused on the following ‘key language-related areas’ in the left hemisphere: Broca’s area, Exner’s area, supplementary motor area (SMA), angular gyrus, Wernicke’s area, and basal temporal language area. For a more detailed analysis, we divided the Broca’s area into two smaller areas (pars triangularis and pars opercularis of the inferior frontal gyrus) and complemented the Wernicke’s area (posterior superior temporal gyrus, pSTG) with the adjacent region in the posterior middle temporal gyrus (pMTG), another key region typically activated by language localizer paradigms (Połczyńska et al., 2017). At each of the three statistical thresholds applied in first-level analyses, we calculated which percentage of the resulting eight ‘key language-related areas’ was activated by the paradigm, using the Brainnetome atlas (Fan et al., 2016) implemented in the ICN_atlas toolbox (Kozák et al., 2017). The respective list of areas from the Brainnetome atlas is presented in Supplementary Table S1. The resulting values were visualized using the seaborn library (https://seaborn.pydata.org/) in Python 3.7.

To test how individual-level activation volume in eight ‘key language-related areas’ was affected by baseline and statistical threshold, a separate repeated-measures ANOVA was conducted for each area. The ANOVAs were conducted on the number of significantly activated voxels in the area and tested the main effects of baseline and statistical threshold and their interaction. Mauchly’s test was used to check the assumption of sphericity. In case it was violated, Greenhouse-Geisser correction was applied. The Bonferroni correction was applied to correct for the number of statistical models, resulting in *α* = .00625.

#### 2.5.4 Test-retest reliability

Consistency of localizer paradigms with regard to individual-level activation is crucial for clinical applications (Binder et al., 2008), so we estimated test-retest reliability of our paradigms. Following Wilson et al. (2017), we used the Dice coefficient (Rombouts et al., 1997) to quantify the similarity of language-related activation in the first and second session of each participant. The Dice coefficient indicated a degree of overlap between the participant’s activation maps in the first and second session and was calculated as follows: Dice = 2*V_overlap_ / (V_1_ + V_2_), where V_1_ and V_2_ denote the number of supra-threshold voxels in the first and second sessions of an individual and Voverlap is the total number of overlapping voxels. The overlap was calculated in Convert3D (http://www.itksnap.org/pmwiki/pmwiki.php?n=Downloads.C3D), which is an extension of the ITK-SNAP tool (Yushkevich et al., 2006). The Dice coefficients can be interpreted as low (.00 to .19), low-moderate (.20 to .39), moderate (.40 to .59), moderate-high (.60 to .79) or high (.80 to 1.00) (Wilson et al., 2017).

The Dice coefficients were calculated for each participant for the frontal, temporal-parietal and frontal-temporal-parietal regions, separately for the SYLL and PW paradigm. Regions were defined using brain lobe masks available in the LI toolbox (Wilke & Lidzba, 2007) based on the atlas by Hammers et al. (2003). The frontal, temporal-parietal and frontal-temporal-parietal regions were selected because they correspond respectively to anterior language regions, posterior language regions and combination thereof, excluding the occipital lobe that is not relevant for the language function. To test what factors affected test-retest reliability, a repeated-measures ANOVA was conducted on Dice coefficients, testing the main effects of brain region, baseline and statistical threshold, as well as all interactions thereof. Mauchly’s test was used to check the assumption of sphericity. In case it was violated, Greenhouse-Geisser correction was applied.

#### 2.5.5 Lateralization indices

Finally, we evaluated the hemispheric lateralization of individual language-related activation. This analysis aimed to confirm the validity of the paradigms and estimate their reliability in establishing individual lateralization of language processing.

Lateralization indices (LIs) were calculated with the LI toolbox for SPM (Wilke & Lidzba, 2007), based on the count and value of suprathreshold voxels and using adaptive thresholding. As implemented in the LI toolbox, adaptive thresholding uses averaged intensity of all voxels in the image as the internal threshold for a given participant, thus taking into account inter-subject variability of BOLD response. LI can take the values from +1 to −1, where +1 stands for full left lateralization of the activation, −1 indicates full right lateralization and 0 indicates bilateral activation. Similarly to Dice coefficients, LIs were calculated for the frontal, temporal-parietal and frontal-temporal-parietal regions for each participant individually, separately for each paradigm in each scanning session.

Given right-handedness of participants in our study, we expected to observe typical left-hemispheric dominance of language-related activation in the majority of participants. This outcome would confirm the validity of the paradigms. Finally, we used a repeated-measures ANOVA to test how LIs were affected by baseline, region, number of session, and interactions thereof. Mauchly’s test was used to check the assumption of sphericity. In case it was violated, Greenhouse-Geisser correction was applied.

## 3. Results

### 3.1 Behavioral results

Participants’ task performance was at ceiling. In the SYLL paradigm, mean sentence completion accuracy was 96.1% (SD 3.1%, range 91.4% – 100.0%) in the first session and 98.4% (SD 1.6%, range 95.0% – 100.0%) in the second session. In the PW paradigm, the mean accuracy was 96.4% (SD 2.4%, range 93.1% – 100.0%) in the first session and 97.5% (SD 1.8%, range 94.9% – 100.0%) in the second session.

### 3.2 Group-level activation maps

For illustration purposes, we demonstrate group-level activation maps from the first session produced at three statistical thresholds: with FWE correction, cluster-size correction and AT (Gorgolewski et al., 2012) (see Figure 2 and Figure 3). A full list of activation clusters comprising more than 100 voxels is presented in Supplementary Table S2 (SYLL paradigm) and Supplementary Table S3 (PW paradigm).

**Figure 2.**
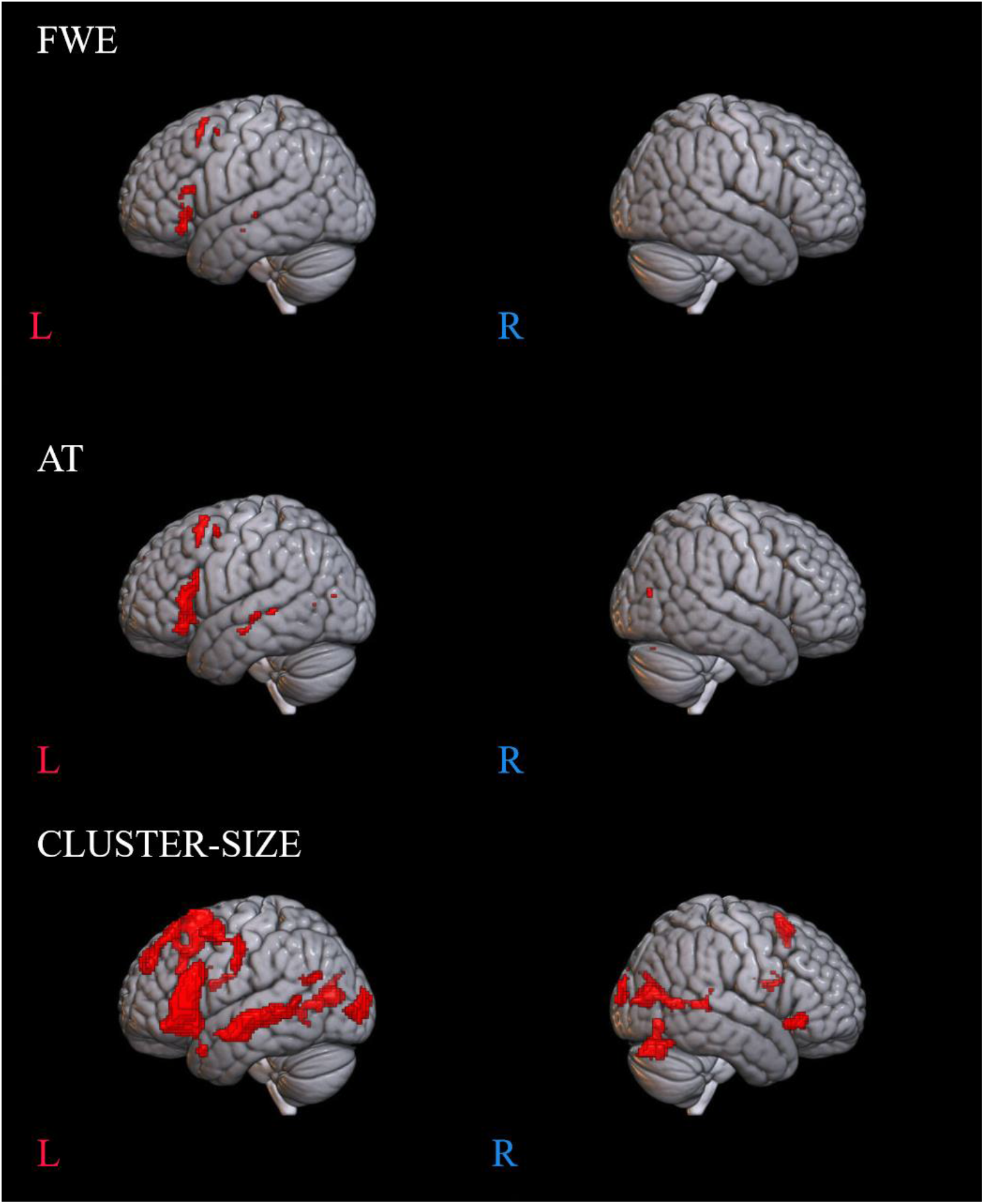
Language-related activation in the SYLL paradigm (paradigm with the syllable baseline). Top: FWE correction for multiple comparisons at *p* < .05, middle: adaptive thresholding (AT) as implemented in Gorgolewski et al. (2012) at *p* < .05, bottom: cluster-size correction for multiple comparisons with a minimum cluster size of *k* ≥ 200 mm^3^ at *p* < .001.

**Figure 3.**
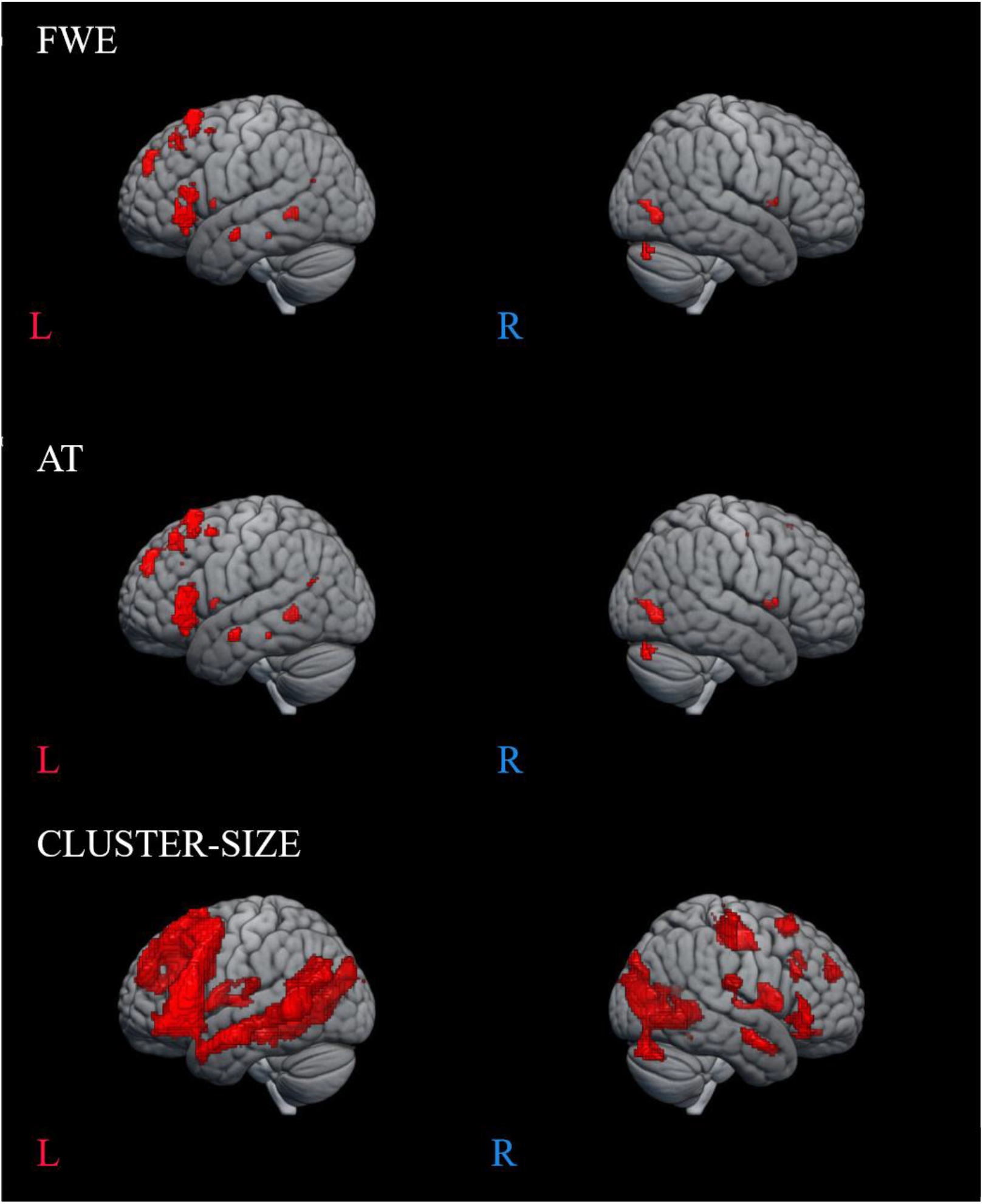
Language-related activation in the PW paradigm (paradigm with the pseudoword baseline). Top: FWE correction for multiple comparisons at *p* < .05, middle: adaptive thresholding (AT) as implemented in Gorgolewski et al. (2012) at *p* < .05, bottom: cluster-size correction for multiple comparisons with a minimum cluster size of *k* ≥ 200 mm^3^ at *p* < .001.

At the most conservative statistical threshold (FWE correction; upper panel in Figures 2 and 3), both versions of the paradigm elicited significant group-level activation in the left inferior and superior frontal gyri. Additionally, the SYLL paradigm activated the orbital gyrus and insula and the PW paradigm activated the left middle and superior temporal gyri. With AT (Gorgolewski et al., 2012; middle panel in Figures 2 and 3), significant group-level activation in both paradigms extended to the left middle frontal gyrus. The activation additionally extended to the left middle and superior temporal gyri in the SYLL paradigm and the orbital gyrus, insula and parts of occipital cortex in the PW paradigm. Finally, when using the most liberal cluster-size correction (bottom panel in Figures 2 and 3), activation in both paradigms extended to a wide network of left frontal, left temporal and bilateral occipital regions, particularly extensive with the PW paradigm.

For illustrative purposes, we also provide individual activation maps from three example participants (first session) in Supplementary Figures S1-S3.

### 3.3 Individual-level activation in key ‘language-related areas’

The violin plots in Figure 4 present the number of activated voxels in ‘key language-related areas’ adopted from Benjamin et al. (2017) across participants (as a percentage of the total number of voxels in the area). The respective numeric values are presented in Supplementary Table S4.

**Figure 4.**
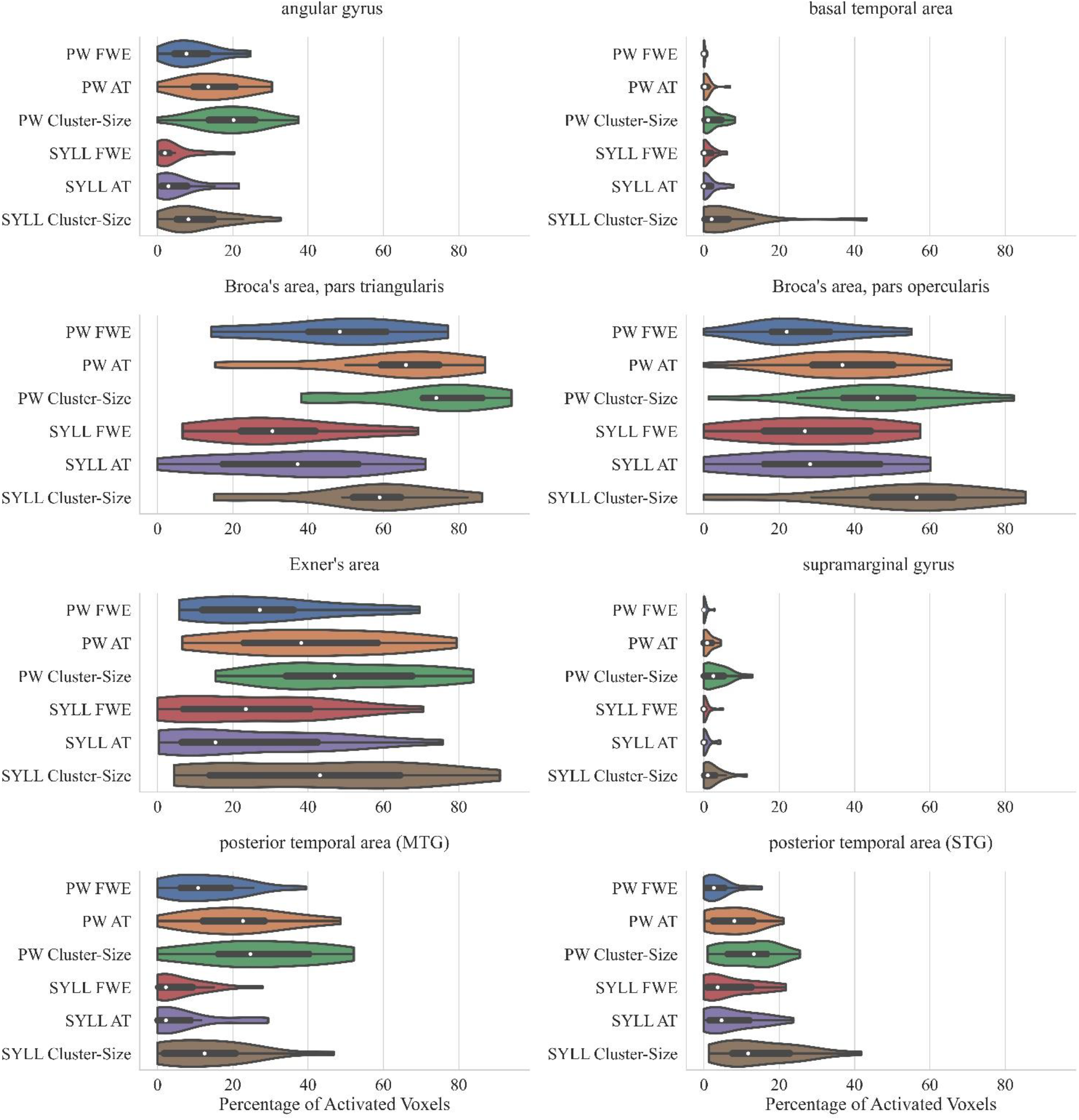
Percentage of significantly activated voxels in ‘key language-related areas’ adopted from Benjamin et al. (2017) depending on the paradigm (PW – pseudoword baseline, SYLL – syllable baseline) and threshold (FWE correction for multiple comparisons at *p* < .05, cluster-size correction with a minimum cluster size of *k* ≥ 200 mm^3^ at *p* < .001, AT: adaptive thresholding as implemented in Gorgolewski et al. (2012) at *p* < .05). The white dot in the middle of each ‘violin’ represents the median value and the thick black bar in the center represents the interquartile range.

As seen in Figure 4, individual-level activation was the most extensive in pars triangularis (median activation ranging from 17% to 59% depending on the paradigm and threshold) and pars opercularis of the inferior frontal gyrus (median activation ranging from 22% to 56%). Individual-level activation was also extensive in the Exner’s area (median activation ranging from 15% to 47%) and the middle temporal gyrus (median activation ranging from 2% to 25%). In other analyzed regions, individual-level activation was less extensive.

Bonferroni-corrected repeated-measures ANOVAs showed that individual-level activation volume was greater with the PW than SYLL baseline in three of ‘key language-related areas’: pars triangularis of the inferior frontal gyrus, *F*(1,17) = 25.22, *p* < .001, posterior middle temporal gyrus, *F*(1,17) = 25.75, *p* < .001, and angular gyrus, *F*(1,17) = 27.97, *p* < .001. In the other five ‘key language-related areas’, individual-level activation volume was not significantly affected by baseline. Expectedly, individual-level activation volume was significantly affected by statistical threshold in six of ‘key language-related areas’: pars opercularis of the inferior frontal gyrus, *F*(1.08, 18.29) = 15.82, *p* = .001, pars triangularis of the inferior frontal gyrus, *F*(1.11, 18.78) = 21.40, *p* < .001, posterior superior temporal gyrus, *F*(1.10, 18.78) = 9.16, *p* = .006, posterior middle temporal gyrus, *F*(1.17, 19.94) = 8.92, *p* = .005, angular gyrus, *F*(1.06, 18.01) = 10.71, *p* = .003, supramarginal gyrus, *F*(1.25, 21.19) = 15.05, *p* < .001. Post-hoc pairwise comparisons showed than in all six regions, cluster-size correction with a minimum cluster size of *k* ≥ 200 mm^3^ at *p* < .001 yielded fewer significantly activated voxels than the FWE correction for multiple comparisons at *p* < .05 (all *p* < .001). No other pairwise comparisons were significant when corrected for multiple comparisons. The interaction between baseline and statistical threshold was not significant in any of ‘key language-related areas’.

### 3.4 Test-retest reliability

Mean Dice coefficients quantifying the overlap of individual activation in the first and second scanning session are presented in Table 1. Mean Dice coefficients ranged from .39 to .61, that is, from low-moderate to moderate-high. A Greenhouse-Geisser-corrected repeated-measures ANOVA demonstrated a significant effect of brain region, *F*(1.04, 17.74) = 6.72, *p* = .018. Post-hoc pairwise comparisons showed that Dice coefficients were higher in the frontal than temporo-parietal (*p* = .019) or frontal-temporo-parietal (*p* = .050) region, and in the frontal-temporo-parietal than temporo-parietal (*p* = .016) region. Dice coefficients were significantly higher with the PW than SYLL baseline, *F*(1, 17) = 5.08, *p* = .038. Finally, there was a significant effect of statistical threshold, *F*(1.41, 23.92) = 6.49, *p* = .011. Post-hoc pairwise comparisons showed that Dice coefficients were higher with the most liberal statistical threshold (cluster-size correction with a minimum cluster size of *k* ≥ 200 mm^3^ at *p* < .001) relative to FWE correction for multiple comparisons at *p* < .05 (*p* < .001) and relative to the adaptive thresholding as implemented in Gorgolewski et al. (2012) at *p* < .05 (*p* = .011). No interactions were significant.

**Table 1.** Dice coefficients in the two paradigms (SYLL: syllable baseline vs. PW: pseudoword baseline) in three regions (frontal, temporal-parietal and frontal-temporal-parietal) at three statistical thresholds (FWE correction for multiple comparisons at *p* < .05, cluster-size correction with a minimum cluster size of *k* ≥ 200 mm^3^ at *p* < .001, AT: adaptive thresholding as implemented in Gorgolewski et al. (2012) at *p* < .05).

| Dice coefficients |  |  |  |  |  |  |
| --- | --- | --- | --- | --- | --- | --- |
| Frontal |  |  |  |  |  |  |
| Baseline | FWE |  | AT |  | Cluster-Size |  |
|  | SYLL | PW | SYLL | PW | SYLL | PW |
| Mean | .49 | .56 | .43 | .59 | .56 | .61 |
| SD | .11 | .12 | .20 | .15 | .09 | .12 |
| Min | .23 | .31 | .02 | .20 | .42 | .34 |
| Max | .67 | .76 | .69 | .78 | .69 | .79 |
| Temporal + Parietal |  |  |  |  |  |  |
| Baseline | FWE |  | AT |  | Cluster-Size |  |
|  | SYLL | PW | SYLL | PW | SYLL | PW |
| Mean | .41 | .47 | .39 | .52 | .54 | .58 |
| SD | .16 | .15 | .21 | .16 | .07 | .10 |
| Min | .14 | .23 | .00 | .15 | .39 | .42 |
| Max | .76 | .71 | .76 | .74 | .65 | .71 |
| Frontal + Temporal + Parietal |  |  |  |  |  |  |
| Baseline | FWE |  | AT |  | Cluster-Size |  |
|  | SYLL | PW | SYLL | PW | SYLL | PW |
| Mean | .49 | .54 | .43 | .56 | .42 | .51 |
| SD | .09 | .11 | .20 | .17 | .15 | .12 |
| Min | .31 | .33 | .01 | .17 | .15 | .26 |
| Max | .66 | .70 | .68 | .74 | .75 | .71 |

The range of Dice coefficients in Table 1, varying between 0 and .79, indicated substantial inter-individual variability. Low Dice coefficients around 0 resulted exclusively following the application of the AT method suggested by Gorgolewski et al. (2012). In some cases, no or almost no voxels survived the statistical threshold established by this method in one of the two participant’s sessions.

### 3.5 Lateralization indices

Table 2 presents mean lateralization indices (LI) for three regions (frontal, temporal-parietal and frontal-temporal-parietal) for the two paradigms (SYLL and PW) in the two scanning sessions. Table 2 also includes Spearman’s correlation coefficients examining reproducibility of the individual LI values across two sessions.

**Table 2.** Lateralization indices for each paradigm (SYLL: syllable baseline vs. PW: pseudoword baseline) in three regions (frontal, temporal-parietal, frontal-temporal-parietal) along with results of Spearman’s correlations between LIs in the two scanning sessions. Significant correlations (*p* < .05) are marked with *.

| Lateralization indices |  |  |  |  |
| --- | --- | --- | --- | --- |
| Frontal |  |  |  |  |
|  | SYLL |  | PW |  |
| Session | Session 1 | Session 2 | Session 1 | Session 2 |
| Mean | .46 | .49 | .50 | .51 |
| SD | .18 | .20 | .16 | .14 |
| Min | .13 | .26 | .25 | .32 |
| Max | .80 | .85 | .88 | .85 |
| Spearman's correlation | $r = .447, p = .063$ | | $r = .496, p = .036^*$ | |
| Temporal + Parietal |  |  |  |  |
|  | SYLL |  | PW |  |
| Session | Session 1 | Session 2 | Session 1 | Session 2 |
| Mean | .33 | .40 | .38 | .32 |
| SD | .17 | .20 | .19 | .16 |
| Min | -.04 | .01 | .10 | .08 |
| Max | .57 | .75 | .88 | .67 |
| Spearman's correlation | $r = .382, p = .117$ | | $r = .603, p = .009^*$ | |
| Frontal + Temporal + Parietal |  |  |  |  |
|  | SYLL |  | PW |  |
| Session | Session 1 | Session 2 | Session 1 | Session 2 |
| Mean | .42 | .46 | .45 | .44 |
| SD | .17 | .18 | .17 | .14 |
| Min | .08 | .20 | .22 | .26 |
| Max | .70 | .84 | .84 | .77 |
| Spearman's correlation | $r = .401, p = .099$ | | $r = .490, p = .038^*$ | |

Mean LIs in all regions across two scanning sessions showed left lateralization regardless of the paradigm (SYLL or PW), ranging from .32 to .51. Individual LIs ranged between very strong left-hemispheric dominance (.88) and bilateral organization (-.04). Minimum individual LIs showing bilateral organization were observed mostly in the temporal-parietal region in both sessions and for both paradigms. The Spearman’s correlation tests showed statistically significant correlation between LIs in the two experimental sessions for the PW paradigm in all regions (all *p* < .05). For the SYLL paradigm, the correlation in regions remained at the level of a statistical trend.

A repeated-measures ANOVA showed a significant three-way interaction between baseline, region and session, *F*(1.11, 18.93) = 8.92, *p* = .006. To address this interaction, separate repeated-measures ANOVAs were performed for the SYLL and PW baseline (there was no main effect of baseline, *F*(1,17) = .06, *p* = .811). With the SYLL baseline, a main effect of region was significant, *F*(1.12, 18.99) = 15.88, *p* = .001, with higher LIs in the frontal than temporo-parietal (*p* = .001) or fronto-temporo-parietal region (*p* = .038) and in the fronto-temporo-parietal than temporo-parietal region (*p* < .001). No other main effects or interactions were significant. With the PW baseline, a two-way interaction of session and region was significant, F(1.08, 18.31) = 6.71, *p* = .017, so separate repeated-measures ANOVAs were performed for the first and second session (there was no main effect of session, *F*(1,17) = .26, *p* = .618). In the first session with the PW baseline, the main effect of region was significant, *F*(1.07, 18.19) = 15.21, *p* = .001. Post-hoc pairwise comparisons showed the same hierarchy of LIs as with the SYLL baseline: LIs were higher in the frontal than temporo-parietal (*p* = .001) and frontal-temporo-parietal region (*p* = .006) and in the frontal-temporo-parietal than temporo-parietal region (*p* = .001). In the second session with the PW baseline, the main effect of region was again significant but much greater, *F*(1.09, 18.52) = 80.49, *p* < .001. Post-hoc pairwise comparisons showed the same hierarchy of LIs as for the first session or for the SYLL baseline: LIs were higher in the frontal than temporo-parietal (*p* < .001) and fronto-temporo-parietal region (*p* < .006) and in the frontal-temporo-parietal than temporo-parietal region (*p* < .001). That is, the significant three-way interaction between baseline, region and session was driven by the main effect of region being the greatest in the second session with the PW baseline.

## 4 Discussion

We presented a new fMRI language localizer for preoperative language mapping in Russian-speaking individuals. Following the world’s best practices (Barnett et al., 2014, Black et al., 2017, Salek et al., 2017, Połczyńska et al., 2017, Unadkat et al., 2019, Wilson et al., 2017, Zacà et al., 2012), the paradigm used a sentence completion task that uniquely engages both language production and comprehension at the word and sentence level. The current study validated the localizer paradigm in a control group of neurologically healthy individuals. In this group, the paradigm successfully activated key language-related areas, elicited expected left-hemispheric lateralization and showed test-retest reliability comparable to previous studies. Apart from demonstrating general validity and reliability of the paradigm, we compared two different baseline conditions (SYLL and PW), for the first time for the sentence completion task. All outcomes were reported at three statistical thresholds.

### 4.1 Activation of key language-related areas

At the group level, both versions of the localizer (with the SYLL and PW baseline) elicited significant activation in an expected network of language-related areas. At the most stringent statistical threshold (FWE correction at *α* = .05), both versions of the paradigm elicited significant activation in the left posterior inferior and posterior superior frontal gyri (differences in results with the SYLL and PW baseline are discussed in Section 4.4). The left posterior inferior frontal gyrus has been implicated in many linguistic processes engaged by the sentence completion task: sentence parsing (Hagoort et al., 2005), conceptual and lexical selection of the completing word (Robinson et al., 2010; Zyryanov et al., 2020), morphosyntactic inflection of the completing word (Den Ouden et al., 2019), and articulatory encoding (Flinker et al., 2007). Such multifaceted involvement of the left inferior frontal gyrus in linguistic processes may explain why its activation was the most statistically robust. The posterior superior frontal gyrus (premotor to supplementary motor cortex) was likely activated as part of the dorsal route supporting articulation (Hickok & Poeppel, 2007) or speech initiation (Dragoy et al., 2020, Kinoshita et al., 2014).

At a more liberal statistical threshold (AT at *α* = .05 as implemented by Gorgolewski et al. (2012)), activation in both versions of the localizer (with the SYLL and PW baseline) extended to the left middle frontal gyrus and to a new cluster of activation encompassing the mid-posterior portions of the left middle temporal gyrus and superior temporal sulcus. More superior and posterior portions of the temporal activation may pertain to phonological processing (Graves et al., 2008, Buchsbaum et al., 2001), whereas activation in the mid part of the middle temporal gyrus may reflect several components of semantic processing, such as storage of heteromodal semantic knowledge (Binder et al., 2009), linkage of word forms to meanings (Bonilha et al., 2017, Hickok & Poeppel, 2007), and semantic control when searching for a completing word (Davey et al., 2016).

Finally, at the most liberal statistical threshold (cluster size correction for multiple comparisons at α = .05), activation extended to a broad left-lateralized frontotemporal and bilateral occipital network. Occipital activation may pertain to reading, including the linkage between visual word processing and phonological word representations (Mano et al., 2013, Richardson et al., 2011): although the baseline condition also required reading, it did not involve any linkage to word representations. Alternatively, the greater occipital activation in the experimental condition may reflect mental imagery of the sentence content (Pearson et al., 2015).

Therefore, group-level results proved that the sentence completion paradigm successfully activated both anterior and posterior areas implicated in language processing. This is in line with previous empirical work (Barnett et al., 2014, Połczyńska et al., 2017, Salek et al., 2017, Unadkat et al., 2019, Wilson et al., 2017, Zacà et al., 2012) and reviews (Black et al., 2017, Manan et al., 2020) promoting sentence-level tasks and particularly sentence completion for eliciting activation in a more comprehensive language network than with word-level tasks. However, significant group-level clusters may result from different individual-level patterns: consistent activation across all/most participants versus strong activation in fewer participants. For clinical use, it is most crucial whether the paradigm consistently elicits significant activation of language-related areas in each tested individual, so that individual maps of language-related areas can be routinely used by a neurosurgeon.

To test this, we analyzed individual-level activation in each participant’s first session. We focused on eight ‘key language-related areas’ adapted with slight modifications from Benjamin et al. (2017). Mirroring the group-level findings, individual-level activation was the most consistent in pars triangularis of the Broca’s area, followed by pars opercularis of the Broca’s area, followed by the Exner’s area. Among these, pars triangularis of the Broca’s area was to some extent activated in each participant (with an exception of the AT statistical thresholding with the SYLL baseline). As represented by the interquartile range of activation area, activation spanned one third to two thirds of this area in most participants. Similarly, most participants showed activation in about one third to one half of pars opercularis of the Broca’s area and, with somewhat greater individual variability, of the Exner’s area. In the vast majority of participants, significant individual-level activation was also present in the posterior middle temporal gyrus, followed by the posterior superior temporal gyrus and the angular gyrus.

On the other hand, most participants did not show significant individual-level activation in the basal temporal area or supramarginal gyrus. The lack of activation in the supramarginal gyrus was surprising, given that activation at the temporo-parietal junction is expected in sentence-level tasks (Połczyńska et al., 2017) and has sometimes also been found with word-level tasks (Roux et al., 2003, Stippich et al., 2007). Still, the majority of participants in the present study showed activation in the adjacent angular gyrus. With regard to the basal temporal language area, it has not been consistently activated by previous fMRI language localizers either (Połczyńska et al., 2017, Barnett et al., 2014, Wilson et al., 2017), despite its long-presumed involvement in lexical retrieval (Krauss et al., 1996, Lüders et al., 1991).

### 4.2 Lateralization of language processing

All participants in the present study were right-handed with no history of neurological disorders. Thus, we expected that the paradigm should elicit primarily left-hemispheric lateralization of language processing, with a certain degree of individual variability (Knecht et al., 2000, Springer et al., 1999). Indeed, mean LI values indicated left-hemispheric lateralization of task-related brain activity, with individual values ranging between bilateral organization and very strong left lateralization. Thus, the ability of the paradigm to detect hemispheric lateralization of language processing activity was confirmed. Numerically, the LI values (mean .32 to .51, depending on the brain region and baseline) were comparable to those in previous studies with neurologically healthy right-handed participants. For example, Deblaere et al. (2002) found individual LI values from –.08 to .58 across four language tasks; Dodoo-Schittko et al. (2012) found mean LI values of .44 and .45 in a verb and antonym generation tasks respectively (for review, see Bradshaw et al., 2017).

Interestingly, language-related activity was significantly more strongly left-lateralized in the frontal (and, correspondingly, frontal-temporal-parietal) than temporal-parietal region. This is in line with contemporary models of language processing. For example, the dual-stream model (Hickok & Poeppel, 2007) postulates a bilaterally organized ventral stream, which primarily involves the temporal lobe and connects speech sounds to meanings, and a left-lateralized dorsal stream, which extends to the frontal lobe and maps speech sounds to articulatory networks. In the same vein, Peelle (2012) argues that phonological and lexical information are processed bilaterally in the temporal lobe, whereas sentence processing engages a left-lateralized pathway including the left inferior frontal gyrus. Although the above models (Hickok & Poeppel, 2007, Peelle, 2012) are mainly concerned with auditory speech processing and differ in the specific division of labor between the frontal and temporal regions, our findings converge with them with regard to stronger left-hemispheric lateralization in the frontal than temporal lobe.

### 4.3 Test-retest reliability

Dice coefficients measuring the spatial overlap of significant activation in the individual’s first and second scanning session were in the moderate range: .39 to .61, depending on the region of interest, baseline condition and statistical threshold. In the only previous study measuring Dice coefficients for the sentence completion task (Wilson et al., 2016), the coefficients indicated a smaller spatial overlap, ranging between .06 and .47 depending on the region of interest and statistical threshold. As a more specific example, at the statistical threshold of α = .001 with the minimum cluster size of 2 cm2, the mean Dice coefficient in the broadest examined region of interest (‘supratentorial region’ in Wilson et al. (2016) and the combination of frontal, temporal and parietal lobe in the present study) was .34 in Wilson et al. (2016) versus .43 or .56 with the SYLL and PW baseline respectively in the present study. The Dice coefficients in the present study were also comparable to those reported in previous studies for other language tasks in neurologically healthy participants, presented in Table 3, and to the mean overlap of .48 across a variety of tasks established in a meta-analysis by Bennett and Miller (2010).

**Table 3.** Comparison of Dice coefficients to previous fMRI paradigms using language tasks in neurologically healthy participants.

| Study | Task | Dice coefficients | Comment |
| --- | --- | --- | --- |
| The present study | Overt sentence completion | .39 to .61 | Group averages depending on the region, baseline and statistical threshold |
| Fesl et al. (2010) | Free reversed association task | .61 | Group average in the global defined language network |
| Wilson et al. (2016) | Picture naming | .38 to .61 | Group averages in the 'supratentorial region', depending on statistical threshold |
|  | Naturalistic comprehension | .30 to .51 |  |
|  | Narrative comprehension | .07 to .37 |  |
|  | Sentence completion | .27 to .47 |  |
| Morrison et al. (2016a) | Phonemic fluency | .36 | Group average, whole-brain |
|  | Rhyming | .54 |  |
| Nettekoven et al. (2018) | Picture naming | .47 or .60 | Group average, depending on statistical threshold |

Test-retest reliability was affected by the brain region and statistical threshold. Dice coefficients were significantly higher in the frontal (and, correspondingly, frontal-temporal-parietal) than temporal-parietal region. This is consistent with higher Dice coefficients in the frontal than temporal or parieto-occipital region of interest in a free reversed association task by Fesl et al. (2010). Conversely, Nettekoven et al. (2018) showed higher Dice coefficients for a picture naming task in the inferior frontal gyrus than the superior temporal gyrus. Possibly, this discrepancy could arise because Nettekoven et al. (2018) used smaller regions of interest than here and in Fesl et al. (2010). With regard to the statistical threshold, Dice coefficients were higher with the most liberal than two more conservative statistical thresholds, in line with previous literature (Nettekoven et al., 2018, Stevens et al., 2013, Wilson et al., 2016).

Hemispheric lateralization of language-related activity also showed moderate test-retest reliability. LIs in the first and second scanning session showed either a significant moderate-to-strong correlation (with the PW baseline) or a statistical trend for a moderate correlation (with the SYLL baseline). This held true when LIs were calculated for the frontal region, temporal-parietal region, and combination thereof. The findings on moderate reliability of the paradigm in identifying both localization and hemispheric lateralization of language-related activity contribute to the literature on general test-retest reliability of fMRI (Bennett & Miller, 2010, Elliott et al., 2020, Holiga et al., 2018).

### 4.4 Comparison of baseline conditions

Each participant was administered two versions of the paradigm with different baselines: reading a sequence consisting of the same syllable and repeating the syllable once more (SYLL baseline) and reading a sequence of pseudowords and repeating any of them once (PW baseline). Both baselines are theoretically plausible for the sentence completion task, yet no previous studies have empirically investigated how their choice may affect the outcomes.

The SYLL and PW baselines showed a very similar spatial distribution of significantly activated areas at the group level (see Section 4.1). Among minor differences in the spatial distribution, one may highlight somewhat more inferior temporal activation with the PW baseline: it largely extended to the inferior temporal sulcus and gyrus, whereas significant activation with the SYLL baseline encompassed more of the superior temporal sulcus and gyrus. Possibly, this pattern emerged because the PW baseline exactly matched the experimental condition in phonological complexity. Therefore, their comparison yielded significant activations in more inferior temporal areas implicated in lexical-semantic processing (Binder et al., 2009, Davey et al., 2016) but not in more superior temporal areas enabling phonological processing (Graves et al., 2008, Buchsbaum et al., 2001).

With regard to the extent of activation, the area of significant group-level activation was somewhat greater with the PW than SYLL baseline across statistical thresholds. This was unexpected: we hypothesized that subtraction of the PW baseline should have yielded a smaller difference from the experimental condition because the PW baseline matched it closer in terms of phonological complexity and real-word neighbors and associations. One possible explanation are differences in how individual participants approached the SYLL baseline: for example, how accurately they tried to pronounce the length (number of vowels) in the string, whether they imposed prosody when reading, et cetera. Possibly, such individual variability could introduce noise in the data, reducing the statistical power of comparison to the experimental condition. Another possible account is that the simpler SYLL baseline allowed more time and cognitive resources for non-task-related cognitive activity, which could also introduce noise in the data.

At the individual level, the paradigms with the SYLL and PW baseline also showed a very similar pattern. With both baselines, activation was most robust across individuals in the inferior frontal gyrus and Exner’s area, followed by the posterior middle and superior temporal gyri and the angular gyrus, whereas the basal temporal area and the supramarginal gyrus showed no activation in most participants (Section 4.1). Mirroring the group-level results, areas of individual activation were numerically lower with the SYLL than PW baseline in most regions of interest. This difference was significant in pars triangularis of the inferior frontal gyrus, posterior middle temporal gyrus, and angular gyrus. With regard to hemispheric lateralization, it did not significantly differ with the SYLL versus PW baseline.

Finally, test-retest reliability, or spatial overlap between significant activation in the participant’s first and second scanning session, was significantly higher with the PW than SYLL baseline. Test-retest reliability of hemispheric lateralization was also higher with the PW baseline. With the PW baseline, the LIs in the first and second session showed a significant moderate-to-high correlation, whereas with the SYLL baseline, they remained at the level of a statistical trend for moderate correlation. This held true for the frontal region, temporal-parietal region and combination thereof.

To summarise, the PW baseline provided more robust activation, as reflected in somewhat more extensive significant activation and higher test-retest reliability. As discussed above, the cognitively simpler SYLL baseline may have allowed more time and cognitive resources for non-task-related cognitive activity or, alternatively, have provoked interindividual variability in the specifics of task performance. Both could introduce noise in the data and reduce statistical power compared to the more cognitively taxing PW baseline. On the other hand, this quantitative difference was not large, and the SYLL baseline appeared to have a qualitative advantage. Namely, posterior temporal activation with the SYLL baseline encompassed more superior areas than with the PW baseline.

Damage to posterior superior temporal gyrus impairs phonological processing (Binder, 2015) and possibly word comprehension, although with some potential for neuroplasticity (Hillis et al., 2017), so its mapping is crucial. Apart from that, the SYLL baseline has the advantage of being cognitively simpler and thus more feasible in the clinical population. Here in the control group of neurologically healthy participants, task performance accuracy was at ceiling and did not differ between the two versions of the paradigm. However, in case of preoperative neuropsychological deficits in patients with brain tumors (Ek et al., 2010, Racine et al., 2015) and epilepsy (Patrikelis et al., 2016), lower cognitive complexity and thus greater feasibility may present an important clinical advantage of the SYLL baseline, despite greater robustness of the PW baseline in the control group.

### 4.5 Comparison of statistical thresholds

We reported all measures and activation maps at three different statistical thresholds. The most conservative was the FWE correction for multiple comparisons at a = .05, followed by the AT method proposed by Gorgolewski et al. (2012) at a = .05, followed by the most liberal cluster-size correction for multiple comparisons with a minimum cluster size of k ≥ 200 mm3 at a = .001. Many previous studies have also reported results at multiple statistical thresholds (Dodoo-Schnitko et al., 2012, Morrison et al., 2016a, Nadkarni et al., 2015, Nettekoven et al., 2018 Wilson et al., 2017), since there is no ‘gold standard’ for statistical thresholding in individual or group-level fMRI analysis. Moreover, studies have shown that the statistical threshold may vastly impact metrics induced from fMRI analysis, such as lateralization indices (Nadkarni et al., 2015) or test-retest reliability metrics (Stevens et al., 2016), so reporting results at only one threshold could be misleading.

In the present study, the spatial distribution of activation both at the group and individual level was expectedly similar across statistical thresholds, although some relevant clusters of activation only emerged at more liberal statistical thresholds. For example, significant group-level activation in the posterior superior temporal gyrus became evident at the two more liberal statistical thresholds, and significant group-level activation in the angular and particularly supramarginal gyrus mainly emerged at the most liberal statistical threshold.

At the individual level, participants highly varied in the extent of activation depending on the statistical threshold: the extent of activation that was present in some participants at the most stringent threshold only appeared in others at more liberal thresholds (Supplementary Table S4). This adds to the evidence for impossibility of using a one-for-all statistical threshold in individual preoperative mapping in clinical practice. Various methods have been proposed in previous literature for individualized statistical thresholding. They have been based, for example, on receiver operating characteristic reliability (Stevens et al., 2016), normalizing statistical maps to the local peak activation amplitude within a brain region (Gross & Binder, 2014, Voyvodic et al., 2009), thresholding based on a fixed percentage of brain activation rather than a statistical threshold (Wilson et al., 2016), and expert judgement by a clinician (American College of Radiology, 2014, Benjamin et al., 2017, 2018). In the present study, we reported the results using one method of individualized thresholding: the AT method by Gorgolewski et al. (2012), which is based on the combination of Gamma-Gaussian mixture modelling with topological FDR thresholding. The AT method did not alleviate individual variability in the extent of activation: the percentage of activation in key language-related areas was not more homogeneous across participants when using the AT method than the two non-adaptive thresholding methods (Figure 4). For clinical practice, this means that the AT method would not solve the issue of largely variable activation strength across individuals that confounds the interpretation of the presence or absence of significant activation in an area. An important research direction, which was beyond the scope of the present study, would be to compare other methods of individualized statistical thresholding.

Test-retest reliability, as measured by Dice coefficients, was in the moderate range across statistical thresholds. Still, Dice coefficients were significantly higher with the most liberal statistical threshold compared to the two more conservative statistical thresholds, in line with previous literature (Nettekoven et al., 2018, Stevens et al., 2013, Wilson et al., 2016). With regard to LIs, these were calculated using adaptive thresholding and taking into account the values of suprathreshold voxels as implemented in the LI Toolbox for SPM (Wilke & Lidzba, 2007), so comparison of different statistical thresholds did not apply to this measure.

### 4.6 Future directions

The present study validated the fMRI language localizer in a control group of neurologically healthy participants. For full validation of the localizer, the crucial next step is to test it in the clinical group of presurgical patients with brain tumors and drug-resistant epilepsy. Data from a clinical sample will test the ability of the localizer to elicit activation in critical language-related areas in patients with different etiology and localization of pathological tissue and thus ultimately assess its clinical value. Data from a clinical sample would also provide the best test case for assessing the clinical value of different methods of individualized statistical thresholding (American College of Radiology, 2014, Benjamin et al., 2017, 2018, Gross & Binder, 2014, Stevens et al., 2016, Voyvodic et al., 2009, Wilson et al., 2016), which remained beyond the scope of the present study.

Finally, as a validation against the gold standard, the findings of the fMRI language localizer in the clinical group will need to be compared to the findings from intraoperative mapping using DES. So far, such comparisons between DES and fMRI language localizer protocols have yielded diverging results (Morrison et al., 2016b, Roux et al., 2003, Spena et al., 2010; for review, see De Witte & Mariën, 2013). Thus, it would be informative to validate our particular fMRI language localizer protocol against DES and thereby add to general evidence on the sensitivity and specificity of fMRI language localizer protocols for preoperative language mapping.

## Data Availability

All data produced in the present study are available upon reasonable request to the authors.

## 6 Author Contributions

KE and SM contributed equally to the manuscript. OD conceptualized, designed and supervised the study. ES and OD created linguistic materials. SM and OB implemented and tested the paradigm. KE, SM, OB and AM collected the data. KE and SM performed data analysis. SM and KE wrote sections of the manuscript. All authors contributed to manuscript revision, read and approved the submitted version

## 8 Acknowledgments

We would like to thank Evgenii Kalenkovich, Olga Buivolova and Valeriya Zelenkova for their help with paradigm development, and all study participants for their contribution.

