## Supplementary Materials for "A new fMRI localizer for preoperative language mapping using a sentence completion task: Validity, choice of baseline condition, and test-retest reliability"

### *Supplementary Material*

**Table S1**. Areas from the Brainnetome atlas (Fan et al., 2016) used to represent the key language-related areas adapted from Benjamin et al. (2016).

| Key language-related area | Areas from the Brainnetome atlas (Fan et al., 2016) |
|---|---|
| Broca's area:<br>(a) IFG, pars triangularis,<br>(b) IFG, pars opercularis | (a) IFG part 4 (rostral area 45), IFG part 3 (caudal area 45), IFG part 2 (inferior frontal sulcus),<br>(b) IFG part 5 (opercular area 44), IFG part1 (dorsal area 44) |
| Exner's area | MFG part2 (inferior frontal junction) |
| SMA | SFG part 4 (dorsolateral area 6), SFG part 5 (medial area 6) |
| Angular gyrus | IPL part 2 (rostrodorsal area 39), IPL part 5 (rostroventral area 39), IPL part 1 (rostrodorsal area 39), IPL part 4 (caudal area 40) |
| Posterior temporal area:<br>(a) pSTG,<br>(b) pMTG | (a) Wernicke's area, pSTG: STG part 4 (caudal area 22), STG part 2 (area 41/42),<br>(b) pMTG: MTG part 1 (caudal area 21), MTG part 3 (dorsolateral area 37) |
| Basal area | Fusiform gyrus part 3 (ventrolateral area 37), Fusiform gyrus part 2 (medioventral area 37) |

*Note*. IFG: inferior frontal gyrus, IPL: inferior parietal lobule, MFG: middle frontal gyrus, pMTG: posterior middle temporal gyrus, pSTG: posterior superior temporal gyrus, SFG: superior frontal gyrus.

**Table S2.** Group-level activation clusters for the SYLL paradigm (paradigm with the syllable baseline), significant at three statistical thresholds (FWE: FWE correction for multiple comparisons at $p < .05$, cluster-size correction with a minimum cluster size of $k \geq 200$ $mm^3$ at $p < .001$, AT – adaptive thresholding as implemented in Gorgolewski et al. (2012) at $p < .05$). Cluster localization was determined in the ICN_atlas toolbox (Kozák et al., 2017) based on the Brainnetome parcellation (Fan et al., 2016). Only clusters of ≥ 100 voxels are presented.

| X | Y | Z | Peak *t*-value | Number of voxels | L/R | Cluster extent |
|---|---|---|---|---|---|---|
| | | | SYLL FWE | | | |
| -28 | 28 | -2 | 10.47 | 442 | L | Inferior frontal gyrus, orbital gyrus, insula |
| -6 | 14 | 60 | 10.7 | 139 | L | Superior frontal gyrus |
| | | | SYLL AT | | | |
| -28 | 28 | -2 | 10.47 | 893 | L | Middle frontal gyrus, inferior frontal gyrus, orbital gyrus, insula, superior frontal gyrus |
| -58 | -24 | -2 | 8.70 | 228 | L | Superior temporal gyrus, middle temporal gyrus, posterior superior temporal sulcus |
| -6 | 14 | 60 | 10.7 | 220 | L | Superior frontal gyrus |
| | | | SYLL Cluster-Size | | | |
| -28 | 28 | -2 | 10.47 | 3037 | L | Middle frontal gyrus, inferior frontal gyrus, orbital gyrus, precentral gyrus, superior temporal gyrus, middle temporal gyrus, insula, basal ganglia |
| 12 | -76 | 12 | 7.63 | 2532 | L | Precuneus, medioventral occipital cortex, lateral occipital cortex, hippocampus; |
| | | | | | R | Precuneus, cingulate gyrus, medioventral occipital cortex, lateral occipital cortex |
| -6 | 14 | 60 | 10.70 | 1886 | L | Superior frontal gyrus, middle frontal gyrus, cingulate gyrus; |
| | | | | | R | Superior frontal gyrus |
| -58 | -24 | -2 | 8.70 | 1671 | L | Superior temporal gyrus, middle temporal gyrus, inferior temporal gyrus, posterior superior temporal sulcus |
| 8 | -72 | -8 | 9.35 | 884 | R | Medioventral occipital cortex |
| -42 | 4 | 56 | 8.83 | 817 | L | Middle frontal gyrus, precentral gyrus, postcentral gyrus |
| -24 | -98 | 10 | 7.45 | 730 | L | Medioventral occipital cortex, lateral occipital cortex |
| 40 | 28 | -6 | 6.88 | 358 | R | Inferior frontal gyrus, orbital gyrus, insula |
| 6 | 10 | 20 | 8.05 | 303 | R | Cingulate gyrus, basal ganglia |
| -48 | -62 | 22 | 6.27 | 225 | L | Posterior superior temporal sulcus, inferior parietal lobule |

*Note.* L: left hemisphere, R: right hemisphere.

**Table S3.** Group-level activation clusters for the PW paradigm (paradigm with the syllable baseline), significant at three statistical thresholds (FWE: FWE correction for multiple comparisons at $p < .05$, cluster-size correction with a minimum cluster size of $k \geq 200$ mm$^3$ at $p < .001$, AT – adaptive thresholding as implemented in Gorgolewski et al. (2012) at $p < .05$). Cluster localization was determined in the ICN_atlas toolbox (Kozák et al., 2017) based on the Brainnetome parcellation (Fan et al., 2016). Only clusters of ≥ 100 voxels are presented.

| X | Y | Z | Peak *t*-value | Number of voxels | L/R | Cluster extent |
|---|---|---|---|---|---|---|
| | | | | PW FWE | | |
| -8 | 50 | 30 | 14.22 | 686 | L | Superior frontal gyrus; |
| | | | | | R | Superior frontal gyrus |
| -52 | 22 | 12 | 13.98 | 673 | L | Inferior frontal gyrus, orbital gyrus, insula |
| 12 | -70 | -2 | 11.71 | 192 | R | Medioventral occipital cortex |
| -14 | -46 | 0 | 9.70 | 123 | L | Parahippocampal gyrus, cingulate gyrus, medioventral occipital cortex |
| -54 | -10 | -12 | 10.22 | 114 | L | Superior temporal gyrus, middle temporal gyrus |
| | | | | PW AT | | |
| -8 | 50 | 30 | 14.22 | 861 | L | Superior frontal gyrus; |
| | | | | | R | Superior frontal gyrus |
| -52 | 22 | 12 | 13.98 | 802 | L | Inferior frontal gyrus, orbital gyrus, insula |
| 12 | -70 | -2 | 11.71 | 253 | R | Medioventral occipital cortex |
| -32 | 16 | 56 | 9.68 | 182 | L | Superior frontal gyrus; middle frontal gyrus |
| -14 | -46 | 0 | 9.70 | 167 | L | Parahippocampal gyrus, cingulate gyrus, medioventral occipital cortex, hippocampus |
| -54 | -10 | -12 | 10.22 | 137 | L | Superior temporal gyrus, middle temporal gyrus |
| | | | | PW Cluster-Size | | |
| -8 | 50 | 30 | 14.22 | 22287 | L | Superior frontal gyrus, middle frontal gyrus, inferior frontal gyrus, orbital gyrus, precentral gyrus, superior temporal gyrus, middle temporal gyrus, inferior temporal gyrus, fusiform gyrus, parahippocampal gyrus, posterior superior temporal sulcus, inferior parietal lobule, precuneus, insula, cingulate gyrus, medioventral occipital cortex, lateral occipital cortex, amygdala, hippocampus, basal ganglia |
| 52 | 28 | 4 | 7.91 | 1212 | R | Inferior frontal gyrus, orbital gyrus, insula |
| -4 | 32 | -14 | 6.71 | 672 | L | Orbital gyrus, cingulate gyrus; |
| | | | | | R | Orbital gyrus |
| 8 | -8 | 52 | 8.42 | 664 | R | Precentral gyrus, postcentral gyrus |
| 38 | -18 | 18 | 7.06 | 631 | R | Inferior parietal lobule, postcentral gyrus, insula |
| -8 | 50 | 30 | 14.22 | 376 | R | Superior frontal gyrus, fusiform gyrus, parahippocampal gyrus, precuneus, cingulate gyrus, medioventral occipital cortex, lateral occipital cortex, hippocampus |
| 54 | -2 | -22 | 7.17 | 352 | R | Superior temporal gyrus, middle temporal gyrus |
| -34 | -22 | 20 | 6.47 | 331 | L | Inferior parietal lobule, postcentral gyrus, insula |
| 14 | 10 | 8 | 10.97 | 213 | L | Basal ganglia, thalamus; |
| | | | | | R | Basal ganglia, thalamus |

*Note.* L: left hemisphere, R: right hemisphere.

**Table S4**. Percentage of activated voxels across participants in key language-related brain regions adopted from Benjamin et al. (2017) out of the total number of voxels in the region based on the Brainnetome atlas parcellation (Fan et al., 2016) depending on the paradigm (SYLL: syllable baseline vs. PW: pseudoword baseline) and statistical thresholding (FWE: family-wise error correction for multiple comparisons at $p < .05$, cluster-size correction with a minimum cluster size of $k \geq 200$ mm$^3$ at $p < .001$, AT: adaptive thresholding as implemented in Gorgolewski et al. (2012) at $p < .05$).

| | ROI | Min | 25th percentile | Median | 75th percentile | Max | IQR |
|---|---|---|---|---|---|---|---|
| PW FWE | Angular | 0.00 | 4.47 | 7.72 | 13.39 | 24.64 | 8.92 |
| | Basal | 0.00 | 0.00 | 0.00 | 0.26 | 0.97 | 0.26 |
| | IFG-tri | 14.31 | 40.10 | 48.43 | 60.69 | 77.19 | 20.59 |
| | IFG-oper | 0.00 | 18.05 | 21.99 | 33.40 | 55.14 | 15.35 |
| | Exner | 5.77 | 11.93 | 27.18 | 36.10 | 69.57 | 24.17 |
| | SMA | 0.00 | 0.00 | 0.00 | 0.00 | 2.85 | 0.00 |
| | pMTG | 0.00 | 6.23 | 10.78 | 19.42 | 39.43 | 13.18 |
| | pSTG | 0.00 | 0.67 | 2.61 | 5.41 | 15.32 | 4.75 |
| PW AT | Angular | 0.00 | 9.63 | 13.54 | 20.69 | 30.37 | 11.06 |
| | Basal | 0.00 | 0.00 | 0.24 | 0.96 | 6.86 | 0.96 |
| | IFG-tri | 15.33 | 59.33 | 65.97 | 74.79 | 86.98 | 15.47 |
| | IFG-oper | 0.00 | 28.76 | 36.78 | 50.28 | 65.79 | 21.52 |
| | Exner | 6.63 | 22.94 | 38.16 | 58.40 | 79.39 | 35.46 |
| | SMA | 0.00 | 0.00 | 0.86 | 2.10 | 4.49 | 2.10 |
| | pMTG | 0.00 | 12.03 | 22.70 | 28.45 | 48.61 | 16.43 |
| | pSTG | 0.09 | 2.36 | 8.07 | 13.17 | 21.19 | 10.80 |
| PW cluster-size | Angular | 0.00 | 13.64 | 20.21 | 25.95 | 37.47 | 12.30 |
| | Basal | 0.00 | 0.24 | 1.13 | 4.63 | 8.32 | 4.39 |
| | IFG-tri | 38.13 | 70.50 | 74.01 | 86.20 | 94.00 | 15.70 |
| | IFG-oper | 1.25 | 37.00 | 46.05 | 55.61 | 82.33 | 18.61 |
| | Exner | 15.46 | 34.14 | 46.99 | 67.64 | 83.93 | 33.50 |
| | SMA | 0.00 | 0.05 | 2.50 | 5.31 | 12.91 | 5.26 |
| | pMTG | 0.00 | 16.18 | 24.68 | 40.21 | 52.15 | 24.03 |
| | pSTG | 1.01 | 6.38 | 13.26 | 16.65 | 25.50 | 10.28 |
| SYLL FWE | Angular | 0.00 | 1.15 | 2.01 | 3.16 | 20.36 | 2.01 |
| | Basal | 0.00 | 0.00 | 0.08 | 1.99 | 6.16 | 1.99 |
| | IFG-tri | 6.65 | 22.16 | 30.52 | 41.97 | 69.25 | 19.81 |
| | IFG-oper | 0.00 | 15.95 | 26.82 | 44.30 | 57.39 | 28.35 |
| | Exner | 0.00 | 6.87 | 23.44 | 40.49 | 70.55 | 33.62 |
| | SMA | 0.00 | 0.00 | 0.00 | 0.07 | 5.06 | 0.07 |
| | pMTG | 0.00 | 0.27 | 2.23 | 9.33 | 27.89 | 9.06 |
| | pSTG | 0.00 | 0.94 | 3.67 | 12.64 | 21.74 | 11.70 |

| | | | | | | | |
|---|---|---|---|---|---|---|---|
| SYLL AT | Angular | 0.00 | 0.99 | 2.91 | 7.87 | 21.63 | 6.88 |
| | Basal | 0.00 | 0.00 | 0.00 | 1.90 | 7.89 | 1.90 |
| | IFG-tri | 0.00 | 17.38 | 37.21 | 53.28 | 71.19 | 35.90 |
| | IFG-oper | 0.00 | 16.07 | 28.20 | 46.77 | 60.15 | 30.70 |
| | Exner | 0.37 | 6.44 | 15.40 | 42.52 | 75.71 | 36.07 |
| | SMA | 0.00 | 0.00 | 0.00 | 0.20 | 4.35 | 0.20 |
| | pMTG | 0.00 | 0.08 | 2.23 | 8.76 | 29.40 | 8.68 |
| | pSTG | 0.00 | 1.47 | 4.68 | 12.02 | 23.76 | 10.55 |
| SYLL cluster-size | Angular | 0.00 | 5.16 | 8.21 | 14.90 | 32.69 | 9.75 |
| | Basal | 0.00 | 0.23 | 2.03 | 6.50 | 43.22 | 6.27 |
| | IFG-tri | 15.05 | 51.92 | 58.96 | 64.61 | 86.15 | 12.70 |
| | IFG-oper | 0.00 | 44.61 | 56.52 | 66.29 | 85.46 | 21.68 |
| | Exner | 4.42 | 13.90 | 43.13 | 64.42 | 90.92 | 50.52 |
| | SMA | 0.00 | 0.11 | 1.03 | 2.94 | 11.41 | 2.84 |
| | pMTG | 0.00 | 1.75 | 12.51 | 20.66 | 46.84 | 18.91 |
| | pSTG | 1.38 | 7.55 | 11.74 | 22.73 | 41.74 | 15.18 |

*Note*. IQR: interquartile range, Angular: angular gyrus, Basal: basal temporal area, SMA: supplementary motor area, Exner: Exner's area, IFG-tri: pars triangularis of the inferior frontal gyrus, IFG-oper: pars opercularis of the inferior frontal gyrus, pSTG: posterior superior temporal gyrus, pMTG: posterior middle temporal gyrus.

**Figure S1.** Individual-level example: Significant language-related activation in the SYLL (panel A) and PW paradigm (panel B) in participant 03BT.

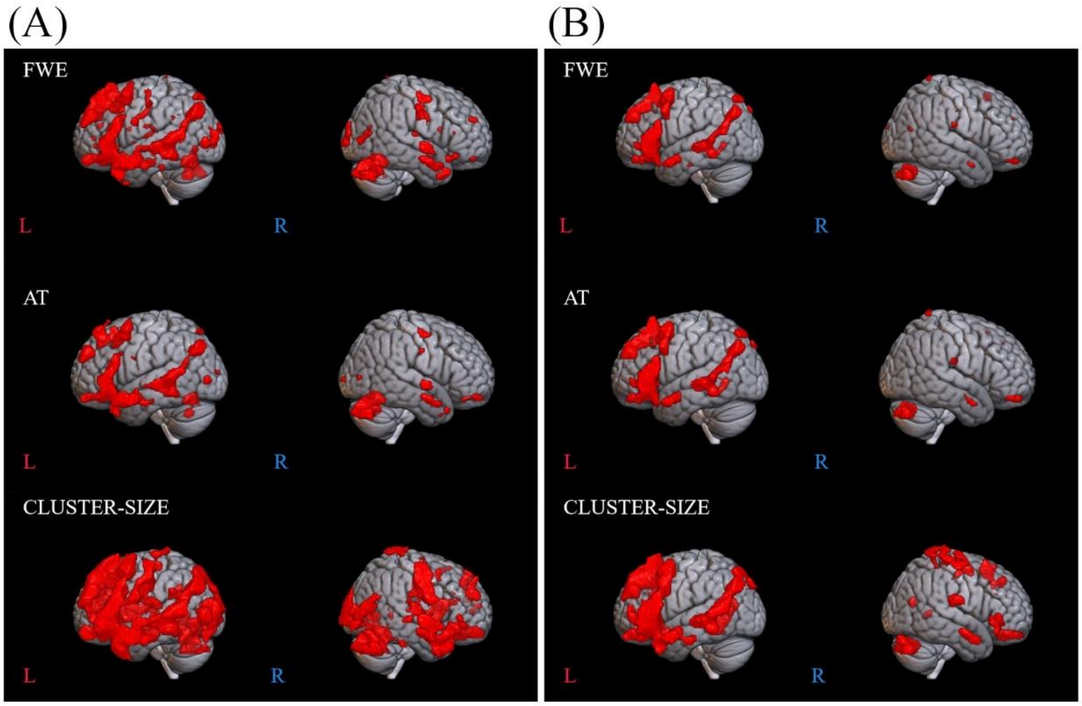


*Note.* Top: FWE correction for multiple comparisons at $p < .05$, middle: adaptive thresholding (AT) as implemented in Gorgolewski et al. (2012) at $p < .05$, bottom: cluster-size correction for multiple comparisons with a minimum cluster size of $k \geq 200$ $mm^3$ at $p < .001$.

**Figure S2.** Individual-level example: Significant language-related activation in the SYLL (panel A) and PW paradigm (panel B) in participant 12UV.

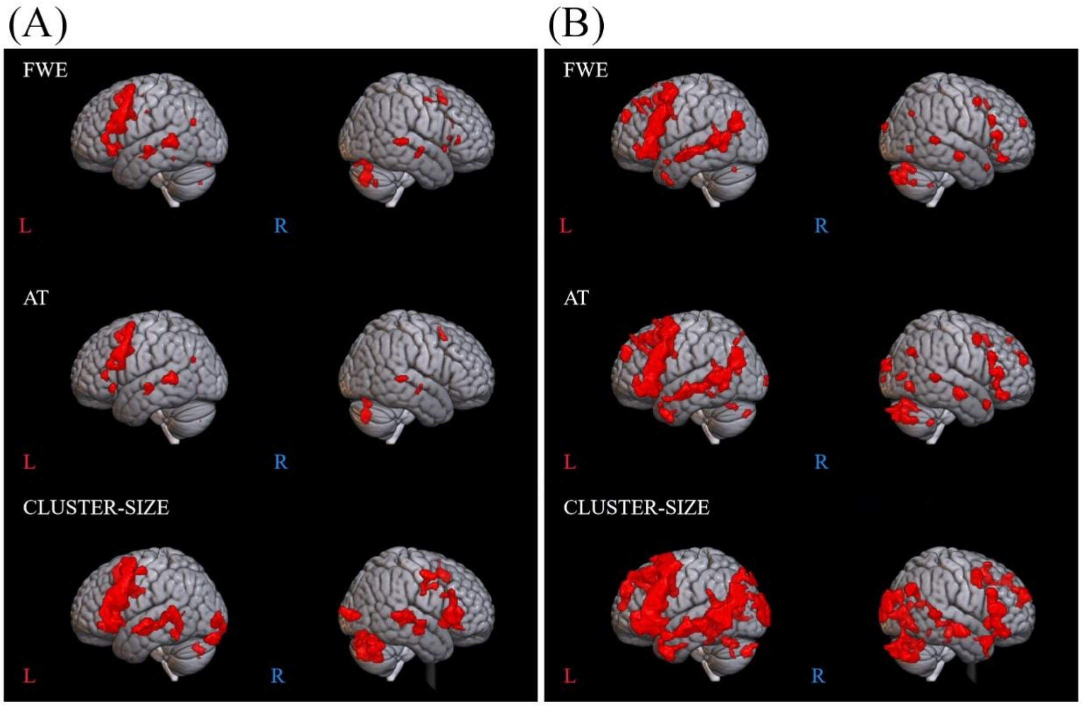


*Note*. Top: FWE correction for multiple comparisons at $p < .05$, middle: adaptive thresholding (AT) as implemented in Gorgolewski et al. (2012) at $p < .05$, bottom: cluster-size correction for multiple comparisons with a minimum cluster size of $k \geq 200$ mm$^3$ at $p < .001$.

**Figure S3.** Individual-level example: Significant language-related activation in the SYLL (panel A) and PW paradigm (panel B) in participant 21BA.

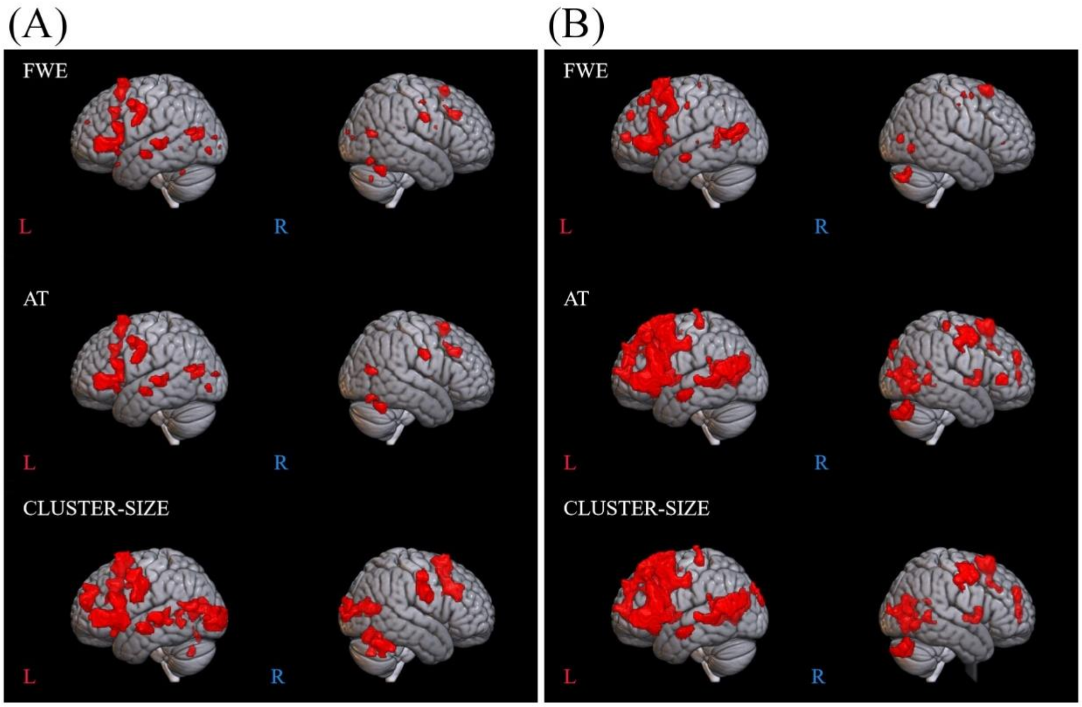


*Note.* Top: FWE correction for multiple comparisons at $p < .05$, middle: adaptive thresholding (AT) as implemented in Gorgolewski et al. (2012) at $p < .05$, bottom: cluster-size correction for multiple comparisons with a minimum cluster size of $k \geq 200$ mm$^3$ at $p < .001$.
